# A Real-World Evaluation of Failure Detection for Liver CT Segmentation

**DOI:** 10.64898/2026.06.26.26356692

**Authors:** Jeddy Bennett, McKell Woodland, Austin Castelo, Mais Altaie, Ajith Anthony, Noreen S. Siddiqi, James P. Long, Kristy K. Brock

**Affiliations:** Department of Imaging Physics, The University of Texas MD Anderson Cancer Center, Houston TX 77030, USA; Department of Mathematics, Brigham Young University, Provo UT 84602, USA; Department of Interventional Radiology, The University of Texas MD Anderson Cancer Center, Houston TX 77030, USA; Department of Biostatistics, The University of Texas MD Anderson Cancer Center, Houston TX 77030, USA

**Keywords:** Out-of-Distribution Detection, Failure Detection, Liver Segmentation

## Abstract

Deep learning models deployed in clinical imaging frequently encounter distribution shifts, yet most out-of-distribution (OOD) detection methods are evaluated only on controlled research datasets. As a result, it is unclear whether existing approaches can reliably identify segmentation failures that arise in real-world clinical practice. We evaluated six OOD detection methods on a deployed liver CT segmentation model (3D nnU-Net) using internal data from 400 patients and external data from 100 patients collected across nearly 70 sites in 7 countries. One method was Pairwise Surface DSC, a surface-based extension of Pairwise DSC, that we introduced. OOD performance was measured using sensitivity, AUROC, and balanced accuracy, with thresholds determined on an independent cohort of 400 patients using the Youden J statistic. Statistical significance was assessed using McNemar tests and stratified bootstraps (*α* = 0.05) with Benjamini–Hochberg correction. Pairwise Surface DSC was the top-performing method, with perfect sensitivities (1.00), near-perfect AUROCs (0.97 internal; 1.00 external), and the highest balanced accuracies (0.94 internal; 0.88 external; *p* < 0.001). These results show that automated failure detection for liver CT segmentation is clinically feasible and that Pairwise Surface DSC is a promising candidate for deployment. Our code is available at https://github.com/mckellwoodland/liver_ct_ood_translation.

## 1 Introduction

Liver cancer is the third leading cause of cancer death globally, with around 765,000 deaths and over 855,000 new cases in 2022 [1]. Effective management of liver disease relies on cross-sectional imaging [2], requiring accurate characterization of liver anatomy for volumetry, surgical planning [3], and lesion analysis. Automated liver segmentation is increasingly used to support these tasks, enabling consistent liver volume estimation [4], reducing the time required for manual annotation [5], and contributing to improved clinical endpoints such as ablative margin assessment [6].

Reliable segmentation is critical for patient safety, yet automated models often struggle with input features that were not present during training [7]. Liver segmentation is particularly vulnerable due to significant shape variability, especially in the presence of hepatic conditions [3]. For example, the only liver CT segmentations deemed clinically unacceptable in Anderson et al. [8] were on scans containing stents and ascites. Out-of-distribution (OOD) detection in safety-critical applications aims to identify such model failures [9].

While OOD detection has been extensively studied in theoretical settings [9], its effectiveness in real-world clinical workflows remains largely unexplored. Contemporary research often relies on curated datasets that fail to capture the complexity of clinical practice, and direct assessments of failure detection are rare [10]. We aim to address these gaps with the following contributions:

1. **Real-world OOD detection evaluation**. We assess failure detection for a deployed liver CT segmentation model using real-world clinical data, including validation on imaging collected from 73 distinct sites (8 countries).
2. **Introduction of Pairwise Surface DSC**. We present a surface-based adaptation of Pairwise DSC [10] that provides a contour-quality measure that is more consistent with clinical acceptability [11].

## 2 Materials and Methods

We conducted a retrospective evaluation of OOD methods for failure detection of a deployed liver CT segmentation model on clinical scans. The study was approved by the The University of Texas MD Anderson Cancer Center Institutional Review Board (PA18-032), with informed consent waived. Our code is available at https://github.com/mckellwoodland/liver_ct_ood_translation.

### 2.1 Data

We queried our internal database for liver-protocol CT studies acquired July to December 2024, resulting in 17,911 scans from 919 patients. We kept scans whose series description specified liver anatomy and removed non-axial, non-diagnostic, and partial-liver scans (1,890 scans remained; 892 patients). 400 unique patients were then separately sampled for validation and testing, selecting one scan per patient. Applying the same criteria to our external database from July to August 2025 yielded 384 scans (100 patients), from which one scan per patient was chosen for external validation. Table 1 summarizes demographics and imaging characteristics, demonstrating clear distribution shifts between the validation and external test cohorts. One scan from an external institution was unintentionally included in the internal test dataset; this affected a single case and did not materially impact the reported results. All imaging was retrieved from UT MD Anderson’s XNAT platform, which ingests and anonymizes PACS data.

**Table 1.** Demographics and imaging attributes, with medians or counts reported. Abbreviations: interquartile range (IQR), manufacturers (mfrs).

| Attribute | Validation | Internal Test | External Test |
| --- | --- | --- | --- |
| Patients | 400 | 400 | 100 |
| Female (%) | 213 (53) | 206 (52) | 42 (42) |
| Age (IQR) | 67 (57-73) | 66 (57-72) | 58 (50-69) |
| Images | 400 | 400 | 100 |
| Voxel Size X/Y (IQR) | 0.8 (0.7-0.9) | 0.78 (0.74-0.82) | 0.78 (0.70-0.87) |
| Slice Thickness (IQR) | 3.0 (2.5-5.0) | 3.0 (2.5-5.0) | 2.0 (1.0-3.0) |
| Sites (Countries) | 3 (1) | 4 (2) | 69 (7) |
| Scanner Models (Mfrs) | 9 (2) | 10 (3) | 31 (6) |

As all scans came from routine clinical workflows, no reference segmentations were available. Three readers assessed model failures: two radiologists (7 and 3 years of experience) and an interventional radiology postdoctoral researcher (3.5 years of experience). For the validation dataset, the radiologist with 7 years of experience labeled segmentations as successes or failures based on whether major edits (edits to ≥ 10% of liver slices) would be required before clinical use. For test datasets, all readers scored segmentations on a five-point Likert scale (Table 2), with scores 1-3 considered failures and 4-5 successes. The final label was determined by majority vote. We reported number of failures, mean Likert ratings, mean pairwise differences between raters, and qualitative observations on failure causes. A two-sided permutation test compared internal and external mean Likert scores (significance level *α* = 0.05).

**Table 2.** Likert scale for determining AI segmentation performance.

| Scale | Description |
| --- | --- |
| 5 Excellent | No edits are required before clinical use. |
| 4 Good | Minor editing is required (i.e., edits on <10% of liver slices). |
| 3 Fair | Major editing is required; edit time < manual contouring. |
| 2 Poor | Major editing is required; edit time > manual contouring. |
| 1 Very Poor | The contour cannot be reasonably edited for clinical use. |

### 2.2 Segmentation

The liver CT segmentation model [6, 12] used in this study was deployed via XNAT before the study began. The model uses the 3D full-resolution nnU-Net framework [13] and was trained on 2,840 scans with five-fold cross-validation on five NVIDIA A100 GPUs (1,000 batches per epoch; roughly 4-5 days). During evaluation, it achieved an average Dice similarity coefficient (DSC) of 0.97 across 248 internal and external scans. Inference was performed using cross-validation ensembling, with all ensemble members sharing the same parameters (patch size [48, 244, 192], target spacing [2.5, 0.98, 0.98]) derived from nnU-Net’s data finger-printing and rule-based configuration. Scans were preprocessed using nnU-Net CTNormalization (clip range [-104,191]) and Z-normalization (mean 63.81, SD 53.44), and postprocessing retained only the largest connected component.

### 2.3 OOD Detection

We evaluated six OOD detection techniques: Maximum Softmax Probability (MSP) [14], Energy Scoring (Energy) [15], Temperature Scaling (Temperature) [16], Deep Ensembling (Ensembling) [17], Pairwise DSC [10], and our proposed Pairwise Surface DSC. For Temperature and Energy, the temperature T was calibrated on the validation dataset over T ∈ {1, 2, 3, 4, 5, 10, 100, 1000}, yielding T=3 for Temperature and T=2 for Energy. For consistency, OOD scores for MSP, Temperature, Pairwise DSC, and Pairwise Surface DSC were inverted (1 - score) so that high values indicated failure. Predictions from the five cross-validation ensemble members were used for Ensembling, Pairwise DSC, and Pairwise Surface DSC. As these predictions were generated during inference, the cost of these methods is limited to lightweight post-processing, making failure detection more practical for clinical deployment than a traditional ensemble.

Pairwise DSC [10] uses the mean DSC across all segmentation pairs for OOD detection. We propose Pairwise Surface DSC, which replaces DSC with surface DSC [18], as surface-based metrics better reflect clinical acceptability [11]. Unlike volumetric DSC, which can remain high despite boundary errors, surface DSC emphasizes boundary agreement by measuring the fraction of surface points within a tolerance (2 mm in our study).

#### Statistical Analyses

OOD detection performance was measured with specificity, sensitivity, balanced accuracy, F1 score, and the threshold-agnostic area under the receiver operating characteristic curve (AUROC). The utilized binary thresholds were determined by maximizing Youden’s J statistic (sensitivity minus false positive rate) [19] on the validation dataset, with failures as the positive class. Nonparametric bootstrapping with 2,000 replicates (400 internal and 100 external scans sampled with replacement) was used to obtain 95% confidence intervals. The significance of performance gains was assessed using McNemar’s test on failures for sensitivity and paired, stratified bootstraps for balanced accuracy and AUROC, using Benjamini-Hochberg (BH)-corrected two-sided *p*-values with significance level *α* = 0.05. Training-based methods (Ensembling, Pairwise DSC, Pairwise Surface DSC) were also compared with output-based methods (MSP, Temperature, Energy) using the same bootstrap tests.

#### Uncertainty Maps

Interpretability is crucial to OOD, allowing model designers and users to identify image regions linked to failure predictions. MSP and Ensembling offer voxel-wise uncertainty estimates that can provide this interpretability, which we demonstrate by overlaying the estimates on a segmented scan.

## 3 Results

### 3.1 Segmentation

The validation, internal test, and external test datasets had 52 (13%), 13 (3%), and 5 (5%) failures, respectively. While the overall failure rates were lower in test data, the rates for the radiologist with seven years of experience remained consistent (10% internal, 12% external). Mean (± SD) Likert ratings of 4.47 ( ±0.42) and 4.49 ( ±0.46) for internal and external data demonstrated no significant difference (*p* = 0.625). The mean pairwise difference between raters was 0.6 Likert points, indicating generally small disagreements.

The most common failure mode was underestimating the left hepatic lobe due to liver hypertrophy, with additional failures in cases with multiple cysts or metastases, portal venous embolization, and distended stomachs. Representative examples are shown in Figure 1, including large cysts (subfigures 1e and 1h), adrenal metastases (subfigure 1f), and extensive metastases (subfigure 1g).

**Fig. 1.**
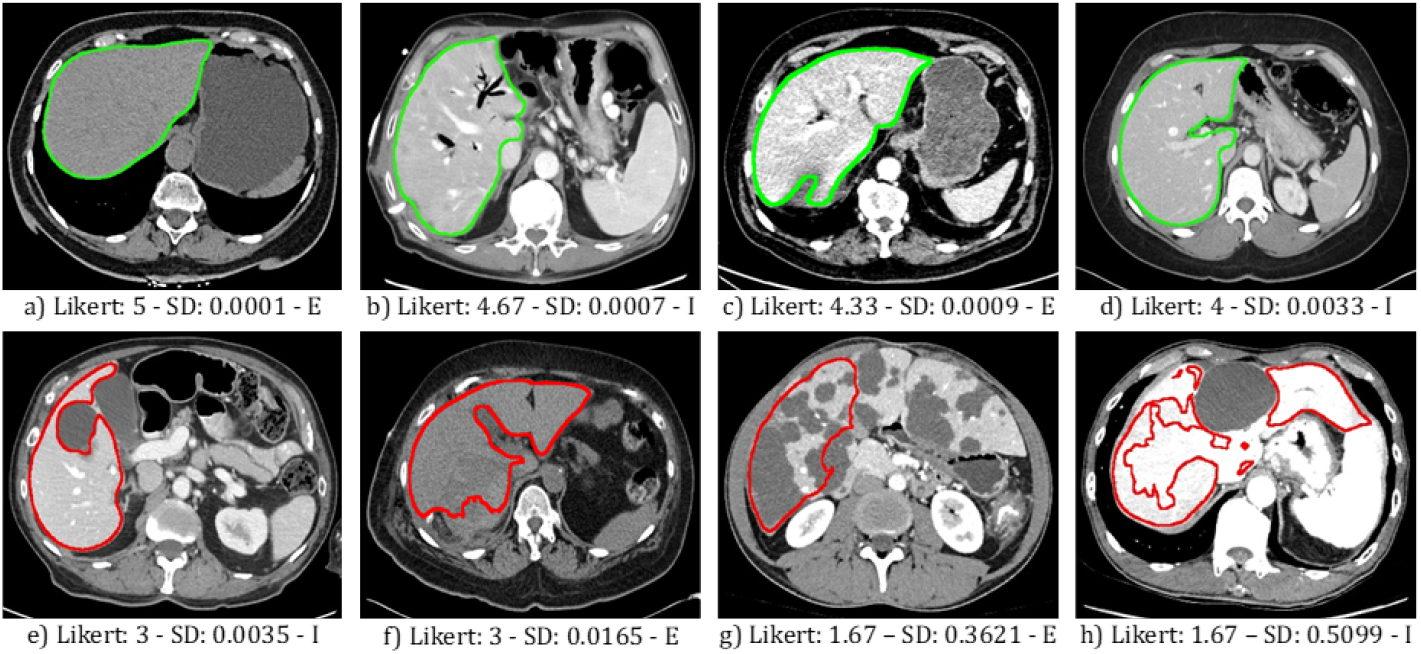
Segmentations grouped by average Likert scores (L) with the corresponding Pairwise Surface DSC (SD). I - internal image; E - external image. Top row: Likert scores 4–5 (green). Bottom row: Likert scores 1–3 (red).

### 3.2 OOD Detection

Table 3 summarizes failure-detection performance on the test datasets, while Figure 2 shows the relationship between Likert ratings and detection scores. Pairwise Surface DSC exhibited the best overall performance. Internally, Pair-wise Surface DSC achieved perfect sensitivity and the highest balanced accuracy, F1 score, and AUROC. Externally, training-based methods performed similarly, with perfect sensitivity, near-perfect AUROC, and high balanced accuracy.

**Table 3.** Performance on internal and external test datasets, with the number of successes (*N*_*S*_ ) and failures (*N*_*F*_ ) reported. Sensitivity (Sens), specificity (Spec), balanced accuracy (Bal Acc), F1 score, and AUROC are reported with 95% confidence intervals. Evaluated methods are MSP, Energy, Temperature (Temp), Ensembling (Ens), Pairwise DSC (P DSC), and Pairwise Surface DSC (PS DSC) . **Bold** values indicate the best performance for each metric. The *†* highlights our proposed method.

| Internal Test Set ($N_S = 387, N_F = 13$) | | | | | |
| --- | --- | --- | --- | --- | --- |
| Method | Sens ↑ | Spec ↑ | Bal Acc ↑ | F1 ↑ | AUROC ↑ |
| MSP | 0.54 (0.25–0.80) | 0.80 (0.76–0.84) | 0.67 (0.52–0.80) | 0.14 (0.05–0.24) | 0.64 (0.45–0.80) |
| Energy | 0.31 (0.07–0.58) | 0.85 (0.81–0.88) | 0.58 (0.45–0.72) | 0.11 (0.02–0.20) | 0.55 (0.35–0.75) |
| Temp | 0.15 (0.00–0.38) | **0.96** (0.94–0.98) | 0.56 (0.48–0.67) | 0.14 (0.00–0.32) | 0.56 (0.36–0.76) |
| Ens | 0.77 (0.50–1.00) | 0.63 (0.58–0.67) | 0.70 (0.56–0.81) | 0.12 (0.05–0.19) | 0.78 (0.59–0.93) |
| P DSC | 0.85 (0.61–1.00) | 0.77 (0.72–0.81) | 0.81 (0.69–0.89) | 0.19 (0.10–0.29) | 0.91 (0.83–0.97) |
| PS DSC† | **1.00** (1.00–1.00) | 0.88 (0.85–0.91) | **0.94** (0.92–0.96) | **0.36** (0.22–0.50) | **0.97** (0.94–0.99) |
| External Test Set ($N_S = 95, N_F = 5$) | | | | | |
| Method | Sens ↑ | Spec ↑ | Bal Acc ↑ | F1 ↑ | AUROC ↑ |
| MSP | 0.40 (0.00–1.00) | 0.52 (0.41–0.62) | 0.46 (0.24–0.76) | 0.08 (0.00–0.19) | 0.60 (0.31–0.95) |
| Energy | 0.00 (0.00–0.00) | 0.60 (0.49–0.70) | 0.30 (0.25–0.35) | 0.00 (0.00–0.00) | 0.37 (0.16–0.56) |
| Temp | 0.20 (0.00–0.67) | **0.83** (0.75–0.90) | 0.52 (0.38–0.79) | 0.09 (0.00–0.29) | 0.47 (0.19–0.81) |
| Ens | **1.00** (1.00–1.00) | 0.54 (0.44–0.64) | 0.77 (0.72–0.82) | 0.19 (0.04–0.33) | 0.97 (0.93–1.00) |
| P DSC | **1.00** (1.00–1.00) | 0.76 (0.66–0.84) | **0.88** (0.83–0.92) | 0.30 (0.08–0.50) | **1.00** (1.00–1.00) |
| PS DSC† | **1.00** (1.00–1.00) | 0.77 (0.68–0.85) | **0.88** (0.84–0.93) | **0.31** (0.08–0.52) | **1.00** (0.98–1.00) |

**Fig. 2.**
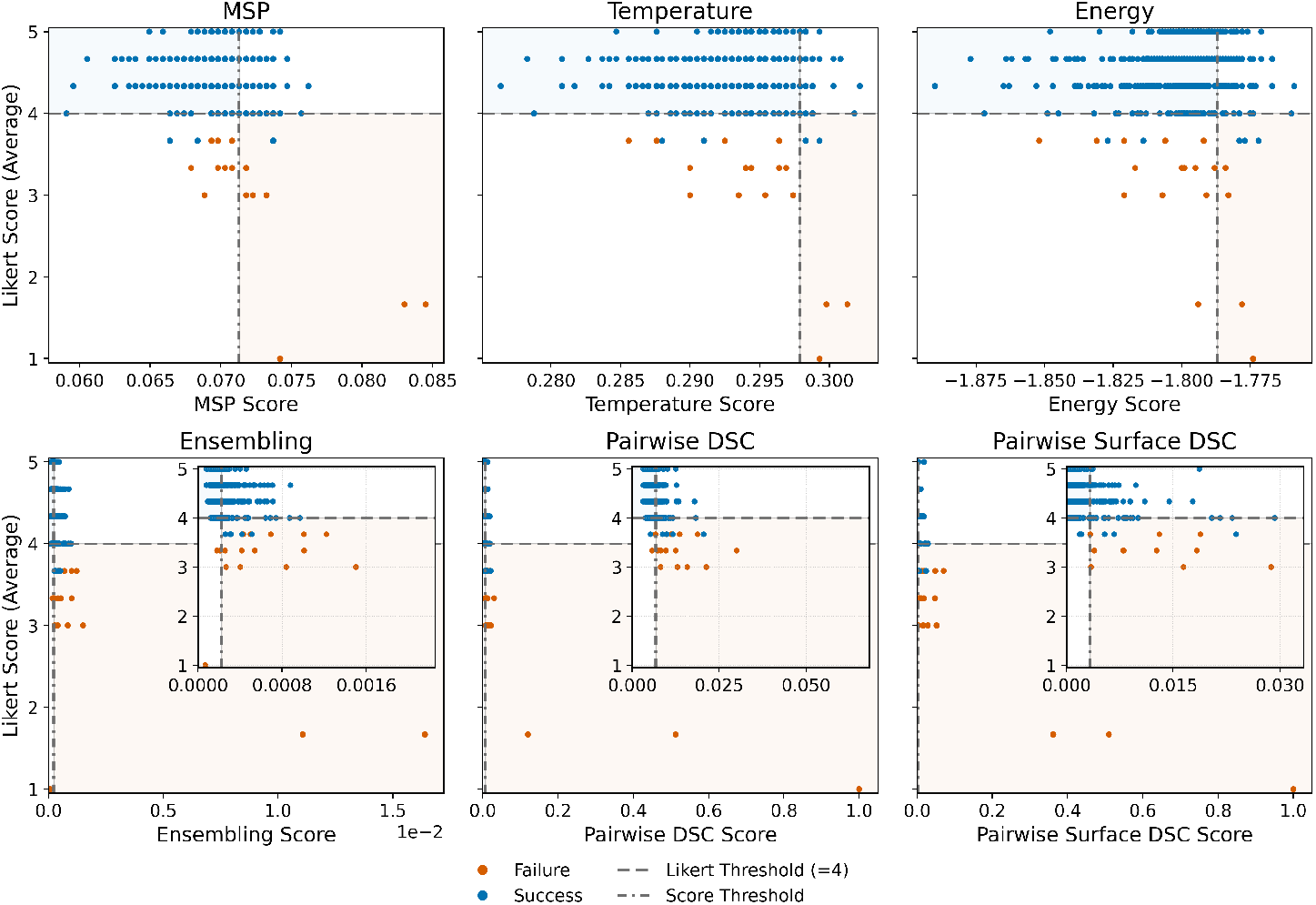
OOD scores versus Likert ratings. The horizontal line marks the success-failure split used before majority voting. Points represent individual scans, colored by true label, with training-based methods shown with an inset zoom near the score thresholds.

#### Statistical Analyses

On the validation dataset, the selected thresholds and their Youden J statistics were: MSP 0.0713 (0.27), Energy -1.7870 (0.14), Tem-erature 0.2979 (0.13), Ensembling 0.0002 (0.64), P DSC 0.0068 (0.59), and Pairwise Surface DSC 0.0033 (0.64). Table 4 reports the *p*-values and statistical powers for all comparisons. Pairwise Surface DSC showed significantly higher balanced accuracy and AUROC than all other methods. For sensitivity, Pair-wise Surface DSC outperformed the output-based methods but not the training-based ones. Overall, the training-based methods had better performances than the output-based methods across all metrics.

**Table 4.** BH-adjusted two-sided *p*-values and power for comparisons of Pairwise Surface DSC (PS DSC) versus each comparator, and for training-vs. output-based method groups, on the combined 500 test scans.

| Comparison | Sensitivity |  | Balanced Accuracy |  | AUROC |  |
| --- | --- | --- | --- | --- | --- | --- |
| | $p$-value | Power | $p$-value | Power | $p$-value | Power |
| PS DSC vs MSP | $p = \mathbf{0.005}$ | 1.00 | $p < \mathbf{0.001}$ | 1.00 | $p < \mathbf{0.001}$ | 1.00 |
| PS DSC vs Energy | $p < \mathbf{0.001}$ | 1.00 | $p < \mathbf{0.001}$ | 1.00 | $p < \mathbf{0.001}$ | 1.00 |
| PS DSC vs Temperature | $p < \mathbf{0.001}$ | 1.00 | $p < \mathbf{0.001}$ | 1.00 | $p < \mathbf{0.001}$ | 1.00 |
| PS DSC vs Ensembling | $p = 0.268$ | 0.00 | $p < \mathbf{0.001}$ | 1.00 | $p = \mathbf{0.011}$ | 0.53 |
| PS DSC vs Pairwise DSC | $p = 0.500$ | 0.00 | $p < \mathbf{0.001}$ | 0.75 | $p = \mathbf{0.032}$ | 0.34 |
| Training vs Output | $p < \mathbf{0.001}$ | 0.97 | $p < \mathbf{0.001}$ | 1.00 | $p < \mathbf{0.001}$ | 0.99 |

#### Uncertainty Maps

Figure 3 shows voxel-wise uncertainty estimates from MSP and Ensembling for the scan in Figure 1g, which failed due to extensive metastases. Both methods highlight uncertainty in areas missed by the segmentation algorithm.

**Fig. 3.**
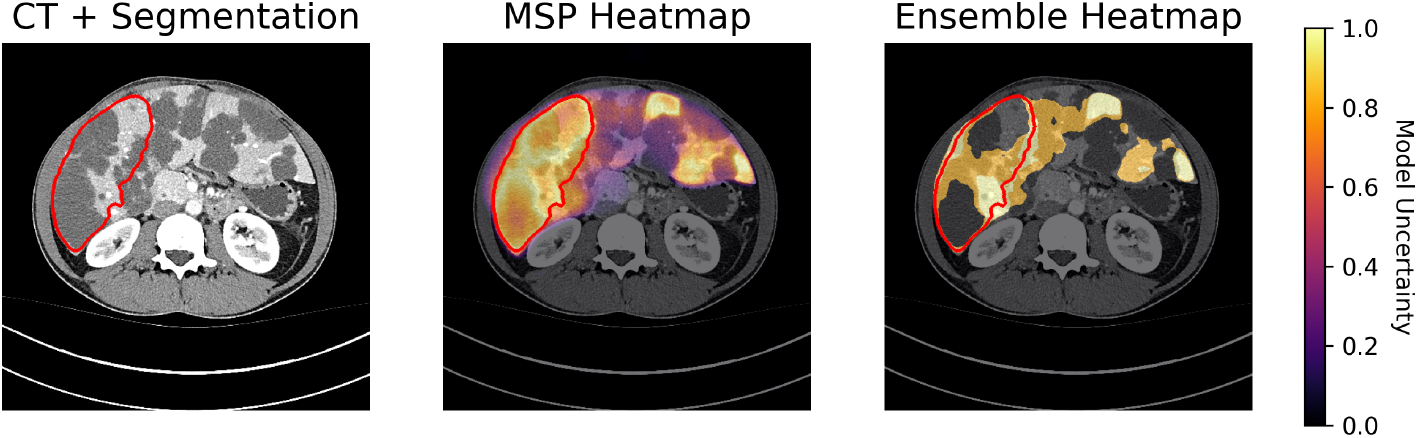
Spatial uncertainty heatmaps for MSP and Ensembling on the slice of Figure 1g show high uncertainty in yellow and low uncertainty in purple/black, with red contours indicating the predicted segmentation.

## 4 Discussion and Conclusion

Most prior work on OOD detection in medical imaging defines OOD through predefined attributes (e.g., controlled perturbations [20], disease types [21], and domain shifts [22]), rarely applying it to the detection of actual model failures, which is the clinically relevant goal. In contrast, our study assesses failure detection for a deployed liver CT segmentation model with clinical data from 73 sites across 8 different countries. We found training-based approaches to be the top performers, consistent with related literature [7, 17]. The improvement of our proposed Pairwise Surface DSC over Pairwise DSC supports evidence that surface-based metrics more effectively indicate clinical acceptability [11].

Calibrating thresholds and evaluating OOD methods in a clinical context requires access to both failures and successes from the target deployment distribution. Identifying failure cases manually is time-consuming and impractical at scale, a challenge exacerbated by the rarity of failures for well-performing models. Furthermore, when a model is updated, threshold performance may decline, necessitating repeated failure identification to recalibrate thresholds. We hope this work motivates further research into threshold selection and OOD evaluation strategies in low-failure settings.

Several aspects of this work’s study design warrant consideration. While expert Likert scoring is more aligned with clinical practice than fixed ground-truth segmentations, it leaves the definition of failure partially subjective. Because the number of failure cases in the test dataset was limited (n=18), statistical power was reported to contextualize each finding. Finally, as this study focuses solely on the liver, future work should evaluate Pairwise Surface DSC on smaller and more complex structures such as tumors.

Our work has multiple practical applications, the most notable being the detection of AI failures in clinical settings, which enhances safety and mitigates automation bias for underrepresented patient groups. Additionally, OOD-based prioritization may aid in model sharing and in enabling large retrospective studies utilizing segmentation by identifying cases needing manual review. Future efforts should adapt these techniques to other organs, modalities, and multiclass scenarios. Overall, our findings indicate that automated failure detection for liver CT segmentation is feasible, and that Pairwise Surface DSC shows promise for deployment.

## Data Availability

The MD Anderson data used in this study may be made available upon request in compliance with institutional IRB requirements and MD Anderson policies and guidelines.

https://github.com/mckellwoodland/liver_ct_ood_translation

## Acknowledgments

Research in this publication was supported by resources of the Image Guided Cancer Therapy Research Program at the University of Texas MD Anderson Cancer Center, the Tumor Measurement Initiative through the MD Anderson Strategic Initiative Development Program (STRIDE), and the National Cancer Institute of the National Institutes of Health under award numbers P30CA016672, 1R01CA221971, R01CA235564, and P01CA261669. N.S.S. is supported by the National Institutes of Health Image-Guided Cancer Therapies T32 Training Program Fellowship Grant (T32CA261856).

